# Novel High-Volume Direct Capture Column for Efficient Nucleic Acid Recovery from Wastewater

**DOI:** 10.64898/2026.07.30.26359326

**Authors:** Lauren M. Aufdembrink, Akli Zarouri, Anil Kumar Meher, Abdennour Abbas

**Affiliations:** PureBioX, Inc., St. Paul, MN, 55114, USA

**Keywords:** Wastewater-based epidemiology (WBE), Nucleic acid extraction, SARS-CoV-2 RNA detection, RT-PCR inhibitors, High-volume processing

## Abstract

The 2020 SARS-CoV-2 pandemic renewed global interest in wastewater-based epidemiology (WBE) as a tool for monitoring public health. Molecular analyses of wastewater are often limited by the small volumes of wastewater that can be processed, due to column clogging, handling constraints, and processing time. Additionally, inhibitors in the complex wastewater matrix reduce the sensitivity of downstream assays such as RT-PCR and sequencing. To address these limitations, we developed a novel column by incorporating a hydrophobic pre-filtration layer and sequential glass fiber filters. This enhanced column design, PureBioX Xpurify Column, enables processing of 1.58× more wastewater (a 58% increase in throughput) while reducing RT-PCR inhibitors and maintaining compatibility with existing workflows. Despite a modest reduction in nucleic acid yield, the modified column consistently improved viral RNA detection sensitivity, including for SARS-CoV-2. This accessible, scalable upgrade strengthens the utility of direct capture methods in WBE-based public health surveillance.

## Introduction

A non-invasive, consistent method for monitoring disease prevalence in communities is critical to public health efforts (1). Wastewater-based epidemiology (WBE) has been employed globally to track drug metabolites, pathogenic microorganisms, and antimicrobial resistance within populations (2–7). The outbreak of the SARS-CoV-2 pandemic in 2020 renewed interest in WBE, emphasizing its value as both an early-warning system and a long-term surveillance strategy for infectious disease outbreaks (4). Numerous pathogens—including Influenza A and B, respiratory syncytial virus (RSV), norovirus, hepatitis viruses, monkeypox, *Salmonella*, and poliovirus—have been or are currently being monitored via WBE, demonstrating its broad applicability in public health surveillance (8–13).

Several techniques are used to concentrate and extract nucleic acids from wastewater, including affinity-based beads, solid-phase extraction, ultracentrifugation, and direct capture via column filtration. Commercially available kits relying on nucleic acid adsorption—such as Promega’s Wizard® Enviro TNA Kit (A2991), Zymo Research’s Quick-RNA Viral Kit (R0135), and Qiagen’s QIAamp Viral RNA Kit (52904)—are commonly employed for WBE. Of these, Promega’s kit has demonstrated the highest SARS-CoV-2 RNA yield using RT-PCR, sequencing, and other molecular techniques (14).

However, these kits are not specifically designed for wastewater processing. They typically require vacuum filtration and centrifugation and can only process up to 40 mL of wastewater, yielding a final total nucleic acid (TNA) sample in 20–40 µL. This limitation is primarily due to two factors: (1) the trade-off between sample volume and the co-extraction of molecular inhibitors, and (2) clogging of direct capture columns by solids and debris in wastewater.

The limited sample volume (maximum 40 mL) significantly restricts the statistical power of WBE. For instance, in the United States, the average wastewater treatment facility processes approximately 34 billion gallons daily (17). A 40 mL sample represents less than 3 × 10^13^ of this total, and samples are often collected only 3–5 times per week due to resource constraints (18). Such limited and infrequent sampling yields high variability and wide confidence intervals, hindering the reliability of pathogen trend analyses and compromising public health decision-making (16).

Another key challenge is the complexity of the wastewater matrix. Current extraction protocols often fail to adequately remove molecular inhibitors—including heavy metals, organic compounds, and microbial metabolites—which negatively affect the sensitivity of downstream assays such as RT-PCR, LAMP, and sequencing (3, 14, 19, 20). While advances are being made in developing inhibitor-tolerant molecular reactions, optimal RNA extraction and purification remain essential for sensitive and accurate pathogen detection (20–24). The nucleic acids recovered from commercial kits often vary in quality and quantity, limiting their utility in sensitive molecular applications.

In this study, experiments were performed aimed at increasing both the volume of wastewater analyzed and the purity of the recovered nucleic acid compared to traditional adsorption-based extraction protocol. Using a custom-built direct capture column with a novel hydrophobic pre-filtration layer stacked above glass filters of decreasing pore size, the column’s capacity was evaluated to improve sample throughput and reduce inhibitory carryover. The custom-built direct capture column was directly compared to Promega’s commercial kit and further comparisons were performed after generation of a novel column with a hydrophobic pre-filter followed by sequential glass fiber filters. These improvements — which are easily integrated into existing systems — enable the processing of over 1.5X volume of wastewater while reducing RT-PCR inhibitors, enhancing the sensitivity of SARS-CoV-2 RNA detection.

## Materials and Methods

### Wastewater Collection

Wastewater was collected as a grab sample at 8:00 AM in a one-gallon plastic Nalgene bottle on the day of each experiment and processed immediately upon receipt. Wastewater was collected by a laboratory technician at the Metropolitan Wastewater Treatment Plant in St. Paul, Minnesota, United States. Samples were collected throughout the year ranging from May 2024 through December 2024.

### Promega Kit Protocol

Processing of wastewater was done following the protocol provided by Promega Corporation (Madison, Wisconsin, United States) for the Promega Wizard® Enviro TNA Kit (catalog number A2991).

### In Laboratory Kit Processing

40 milliliters (mL) of wastewater was centrifuged at 3000 RPM on an Eppendorf Centrifuge 5810 R for ten minutes. Supernatant was poured off and the pellet was discarded. 13.5 mL of lysis buffer was added to supernatant. Lysis Buffer is 6 M guanidine thiocyanate (Chem-Impex International Inc., Wood Dale, Illinois) and 10% polyethylene glycol 20000 (Sigma-Aldrich). Samples were mixed by inverting and incubated at room temperature for 30 minutes. 48 mL of isopropanol was added to sample and mixed by inverting. Samples were processed through direct capture column with vacuum filtration (50-80 Hg). Direct capture columns were made using 4 levels of silica stacked with pore sizes of 1.2 µm, 4 layers of glass (Cytvia Life Sciences, Catalog #DNA050B-8) with pore sizes of 1.2 µm or 4 layers of glass (Cytvia Life Sciences, Catalog #DNA050B-8) with pore sizes of 1.2 µm and a single 50 µm hydrophobic layer (Shenzhen Baimai Life Science Co. Ltd) from bottom to top of filter, respectively, depending on the experiment being conducted. Column was washed with 5 mL of Wash 1. Wash 1 is 3.5 M guanidine hydrochloride (Chem-Impex International Inc., Wood Dale,Illinois) and 40% isopropanol. Column was washed with 20 mL Wash 2. Wash 2 is 2 mM Tris-hydrochloride (Sigma-Aldrich), 20 mM Sodium Chloride (Sigma-Aldrich) and 80% ethanol, pH 7.5. Direct capture column was taken off vacuum and put in a 25 mL conical tube. 500 µl of releasing buffer was applied to column. Releasing buffer is 8 M guanidine hydrochloride. Column was spun at 3000 RPM for 1 minute. 2X 500 µl water was applied to column and then spun at 3000 RPM for 1 minute. The 1500 µl of elution was swirled to mix and 1500 µl of isopropanol was added. Tube was swirled to mix well. Elution mixture was applied to a mini-spin column (catalog number: DNA050B-8, Shenzhen Baimai Life Science Co. Ltd., sourced via Alibaba)) with 1 min spins at 3000 RPM. Mini-column was washed with 300 µl wash 1, 2x 500 µl wash 2 and eluted with 2x 20 µl water using 1 min 3000 RPM spins resulting in 40 µl sample of extracted nucleic acids.

### Total Nucleic Acid Readings

Samples were at room temperature and measured on the Nanodrop One (ThermoFisher Scientific) using the RNA selection.

### Reverse Transcriptase Real Time PCR

Primers and probes were ordered premixed from Integrated DNA Technologies (Des Moines, Iowa). nCOV_N1 Forward Primer: catalog number 10006821, nCOV_N1 Reverse Primer: catalog number 10006822, nCOV_N1 (FAM) Probe: catalog number 10006823, nCOV_N2 Forward Primer: catalog number 10006824, nCOV_N2 Reverse Primer: catalog number 10006825, nCOV_N2 (SUN) Probe: catalog number 10007050. The primer probe sets were chosen due to the recommended use by the Center for Disease Control (25). For the RT-PCR, 5 µl (unless normalizing nanograms in reaction) of eluted nucleic acid was added to 15 µl of master mix. Master mix was 0.5 µm N1 forward primer, 0.5 µm N1 reverse primer, 0.5 µm N2 forward primer, 0.5 µm N2 reverse primer, 0.12 µm N1 FAM probe, 0.12 µm N2 SUN probe, 5 µl TaqMan™ Fast Virus 1-Step Multiplex Master Mix for qPCR (No ROX) (ThermoFisher Scientific, catalog number: 5555536), nuclease free water up to 20 µl. Program: 25 °C for 2 minutes, 50 °C for 15 minutes, 95 °C for 2 minutes, 45 cycles of 95 °C for 3 seconds, 55 °C for 30 seconds. Reactions were run on the QuantStudio™ 3 Real-Time PCR System (Applied Biosystems by ThermoFisher Scientific). Runs analyzed and threshold values were determined using the QuantStudio™ Design & Analysis Software v1.5.2 (Applied Biosystems by ThermoFisher Scientific).

### Method Comparison

TNA concentration and cycle threshold of multiplexed real-time reverse transcription PCR (RT-PCR) detecting SARS-CoV-2 was compared. Samples were compared on a side to side basis and not across multiple days.

## Results

### Direct Capture Column Matrix

Previous studies have demonstrated the potential of non-conventional column matrices—materials other than silica gel—for improving nucleic acid extraction quality (26). To evaluate this in the context of wastewater-based RNA recovery, we compared a column stacked with commercially available silica to one containing Cytiva Whatman™ Grade GF/B glass microfiber filters. According to Cytiva’s internal data, these filters exhibit enhanced binding capacity for cell-free DNA and RNA fragments (<50 bp) compared to silica-based columns. Additionally, when tested with 10 kilobase (kb) plasmid DNA, the Cytiva glass fiber-based filters achieved greater recovery efficiency than silica columns.

Given the broad size range of RNA viral genomes—ranging from approximately 1.8 kb (e.g., *Saccharomyces cerevisiae* killer virus M1) to 33.5 kb (e.g., *Ball python nidovirus*)—the Cytiva filters were selected to optimize recovery of longer viral RNAs, such as SARS-CoV-2 (29.8 kb) (27, 28).

Use of the Cytiva glass fiber-based filters led to a statistically significant (p < 0.05) 108.13% increase in total nucleic acid (TNA) yield, from 67.8 ng/µL to 141.11 ng/µL, without altering the elution volume. Moreover, SARS-CoV-2 detection by RT-PCR showed a reduction in cycle threshold (Ct) values, indicating improved sensitivity (Figure 1). Based on these findings, all subsequent experiments were conducted using the Cytiva glass filter-based column configuration.

**Figure 1.**
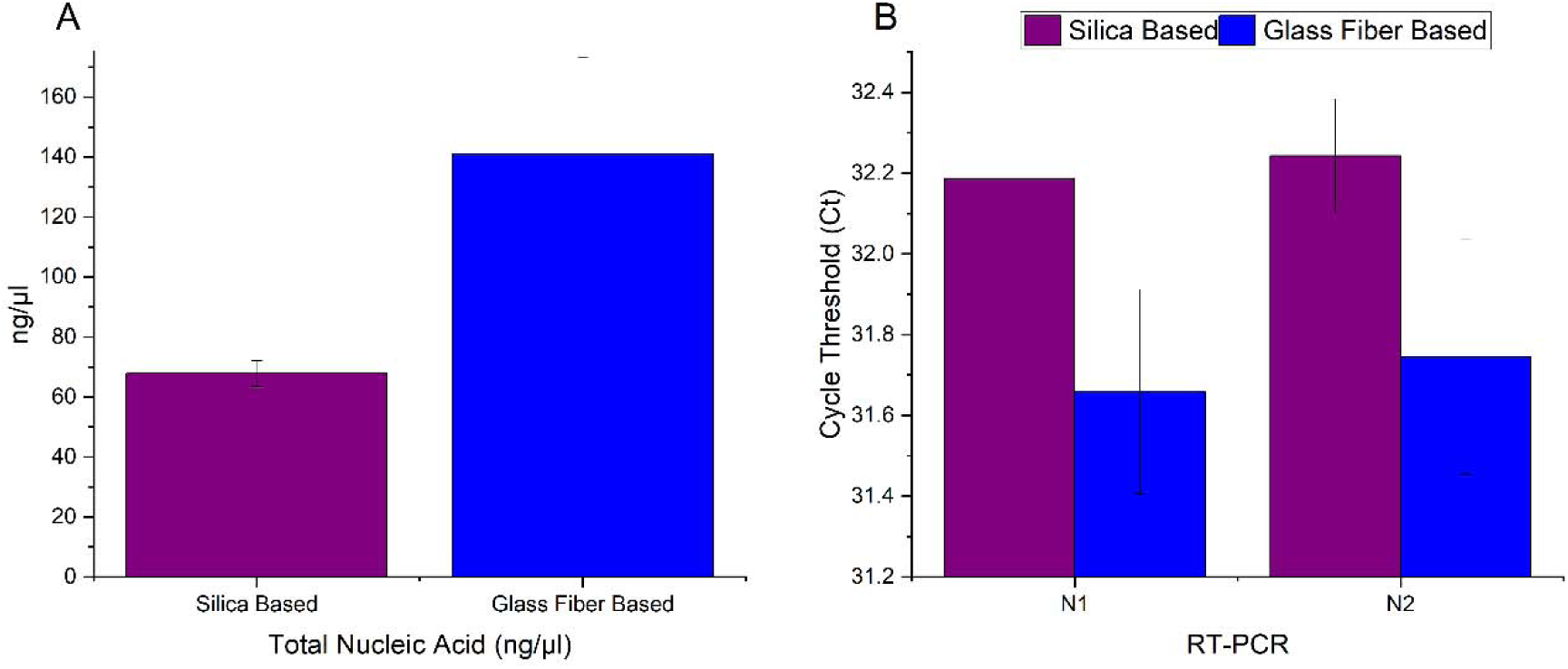
Comparison of nucleic acid extraction efficiency between commercially available silica filters and Cytiva glass fiber filters used in large direct capture columns. Wastewater samples were processed using columns stacked with either silica filters (purple bars, *n* = 3) or Cytiva glass fiber-based filters (blue bars, *n* = 3). **(A)** Cytiva filters yielded more than twice the total nucleic acid compared to silica filters (*p* = 0.017). **(B)** Nucleic acid extracted with Cytiva filters resulted in lower cycle threshold (Ct) values for both SARS-CoV-2 targets (N1 and N2), indicating enhanced detection sensitivity (N1: *p* = 0.066; N2: *p* = 0.056)

### Increasing Volume Capacity

To enable the processing of larger wastewater volumes than those supported by traditional extraction kits, a novel direct capture column incorporating a stacked filtration design was developed. Conventional nucleic acid-binding columns typically utilize materials with small pore sizes—as small as 60 Å—which are effective for nucleic acid separation via chromatography (29). However, such small pore sizes are prone to rapid clogging when processing complex matrices like wastewater, which contains high levels of solids and debris.

To address this limitation, a column design was implemented in which pore size gradually decreased through sequential layers. A hydrophobic pre-filter (HP) with a larger pore size was placed above four layers of the Cytiva glass microfiber filters, forming a high-throughput “HP stack” column (Figure 2). HP filters with pore sizes ranging from 20 µm to 100 µm were evaluated, with 50 µm identified as the optimal configuration based on flow and performance characteristics (Supplemental Figure 1). This column is referred to as PureBioX HP Glass Stack.

**Figure 2.**
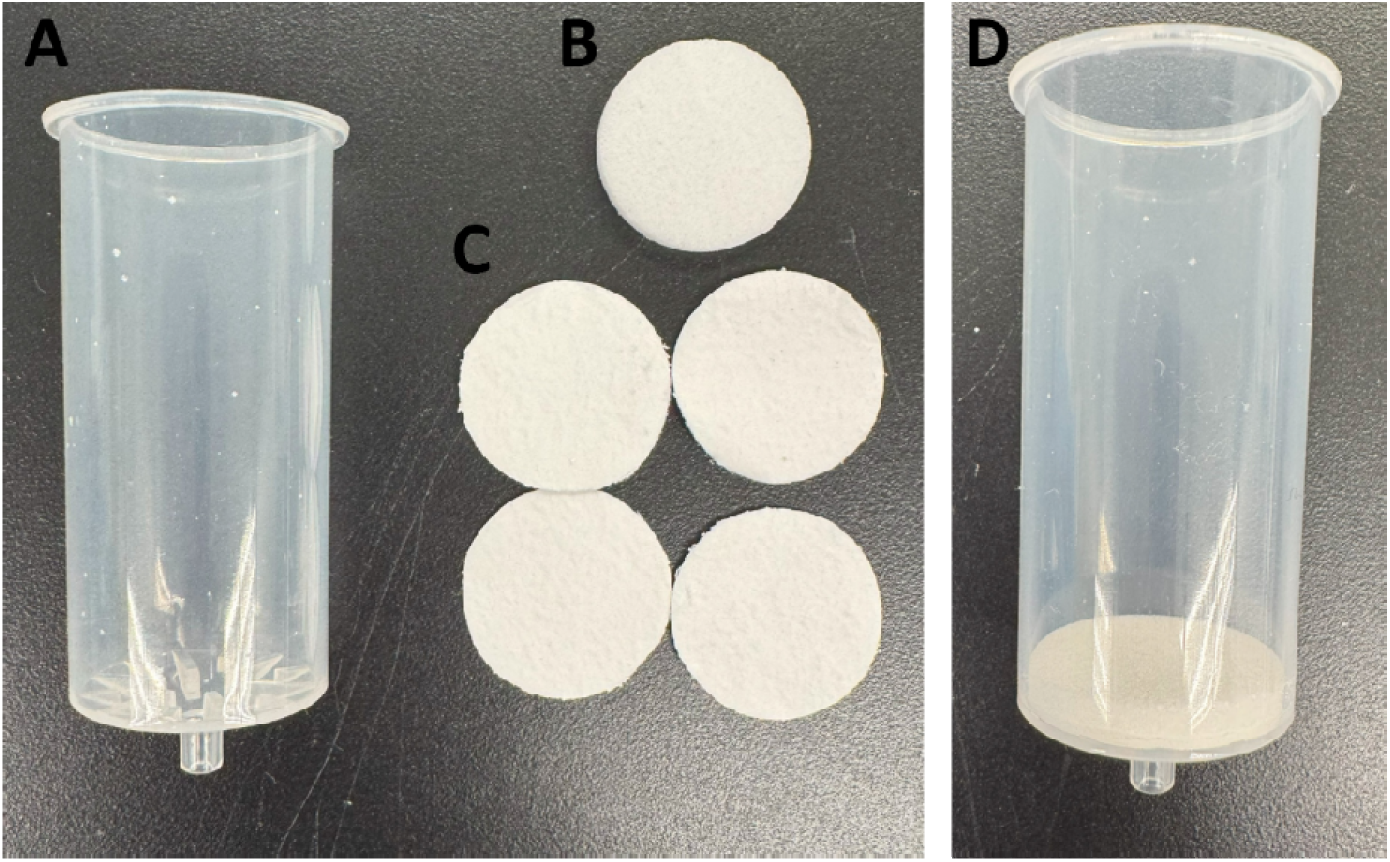
PureBioX HP Glass Stack Column, (A) Plastic cylinder (B) Hydrophobic filter later (C) 4 layers of Cytiva glass fiber filter. (D) Assembled PureBioX Xpurify Direct Capture Column V1

The PureBioX HP Glass Stack column significantly increased the volume of wastewater that could be processed without clogging by 56.67% (48.91 versus 76.63 mL), compared to columns with glass filters alone (Figure 3A). While TNA yield was reduced with the PureBioX HP Glass Stack column addition (1228.57 versus 716.97 ng/µL) (Figure 3B), there was no significant difference in SARS-CoV-2 detection (N1: *p* = 0.107; N2: *p* = 0.860), as reflected in comparable RT-PCR Ct values (Figure 3C). This suggests that the HP layer may facilitate more selective enrichment of target viral RNA or improve removal of inhibitory substances, thereby preserving analytical sensitivity despite lower total nucleic acid recovery.

**Figure 3.**
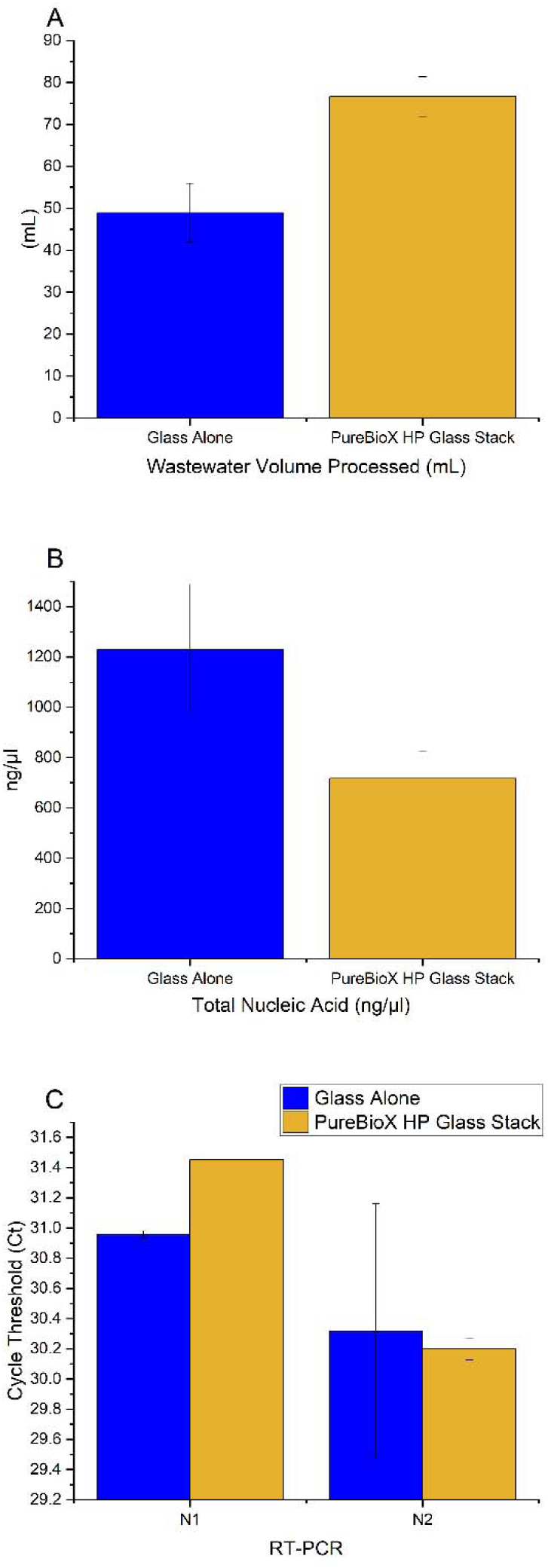
Comparison of nucleic acid extraction performance between columns containing only Cytiva glass filters (blue bars, *n* = 2) and the novel PureBioX HP Glass Stack column, which incorporates a hydrophobic pre-filter above the glass layers (yellow bars, *n* = 2). (A) The PureBioX HP Glass Stack processed 56.67% more wastewater in 10 minutes than the glass-only column (*p* = 0.044). (B) Total nucleic acid (TNA) yield (ng/µL) was lower for the PureBioX HP Glass Stack compared to glass alone, though not statistically significant (*p* = 0.123). (C) Ct values for SARS-CoV-2 detection (N1 and N2 targets) were comparable between the two column types (N1: *p* = 0.107; N2: *p* = 0.860), indicating preserved detection sensitivity despite the lower TNA yield.

### Comparison to Promega

With the increase volume capability compared to traditional columns, the PureBioX HP Glass Stack column was tested, using lab-made produced lysing and washing buffers, generating the PureBioX Xpurify Kit, against the commercially available Promega Wizard® Enviro TNA Kit. 100 mL of wastewater was processed with the PureBioX Xpurify Kit, more than doubling the conventional volume of 40 mL that Promega’s kit processes in the same amount of time (10 mins). Testing wastewater collected on 3 different days, with the PureBioX Xpurify Kit and Promega method side by side, showcased that an increase in TNA yield (570.14 versus 292.41 ng/µL, *p* = 0.011) and a decrease in Ct values occurred with the higher volume processed (Figure 4A&B). This was expected due to the larger volume of wastewater processed.

**Figure 4.**
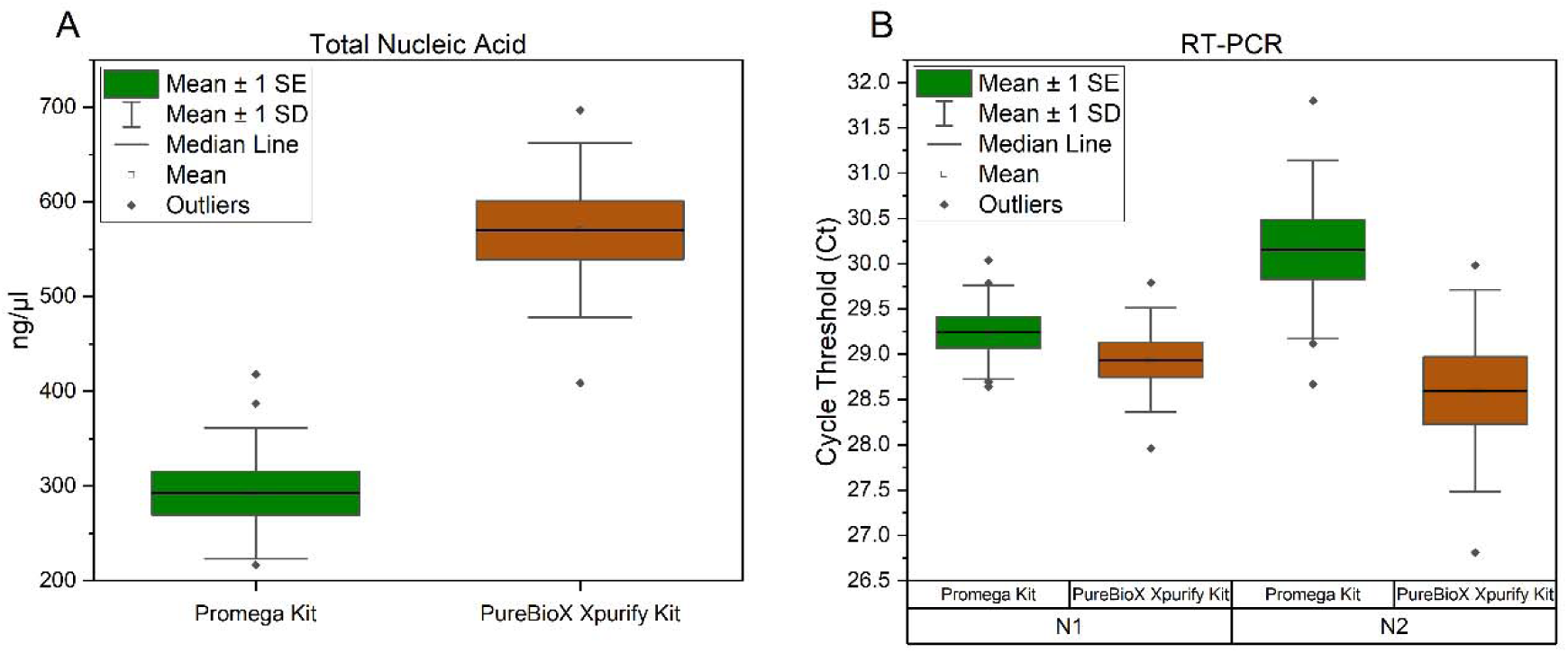
Comparison of nucleic acid extraction performance between the PureBioX Xpurify Kit using the HP-stacked column (Orange) and the Promega Wizard® Enviro TNA Kit (green bars). Wastewater samples were processed in triplicate on the day of collection across three separate days. Replicates were averaged for each day and plotted. (A) TNA yield (ng/µL) was significantly higher using the PureBioX Xpurify Kit with the PureBioX HP Glass Stack column (*p* = 0.011). (B) Ct values from RT-PCR targeting two regions of the SARS-CoV-2 nucleocapsid gene: N1 (left, *p* = 0.499) and N2 (right, *p* = 0.083). Both targets showed decreased Ct values with the PureBioX Xpurify Kit, however, not statistically significant.

However, there were issues with applying the PureBioX Xpurify Kit Lysis buffer for reliable use. The Lysis buffer (6 M guanidine thiocyanate, 10% PEG 20K) was not fully dissolved at room temperature; it had to be heated to 60 °C to for the salts to completely go into solution. If not properly heated, the buffer would crash out of solution, altering the amount of chaotropic salts in solution leading to varying lysis conditions from sample to sample rendering the results unreliable or comparable. Even with proper heating, the buffer would often leave debris on the measuring devices, indicating salts not making it into the lysis process. This poses a problem when trying to create an easy to use, consistent method. The concentration of chaotropic salts plays a pivotal role in lysing viral capsids and adsorption of nucleic acid to the direct capture columns (30,31). A literature search for buffers used on nucleic extraction in wastewater was completed but none were found to be completely soluble at room temperature, and achieve the concentration of chaotropic salt desired for proper lysis and nucleic acid binding (Supplemental Figure 2) (32–35).

### Development of the PureBioX Xpurify Column

To integrate the consistency of the Promega lysis buffer with the enhanced volume-handling capabilities of the HP-stacked design, a novel column figuration was generated by incorporating a hydrophobic (HP) filter layer directly above the Promega column’s direct capture matrix, as in the PureBioX HP Glass Stack Column. This novel column is referred to as the PureBioX Xpurify Column.

Figure 5 illustrates the physical differences between the standard Promega column and the PureBioX Xpurify Column, highlighting the addition of the HP filter layer that enables improved wastewater flow and volume capacity.

**Figure 5.**
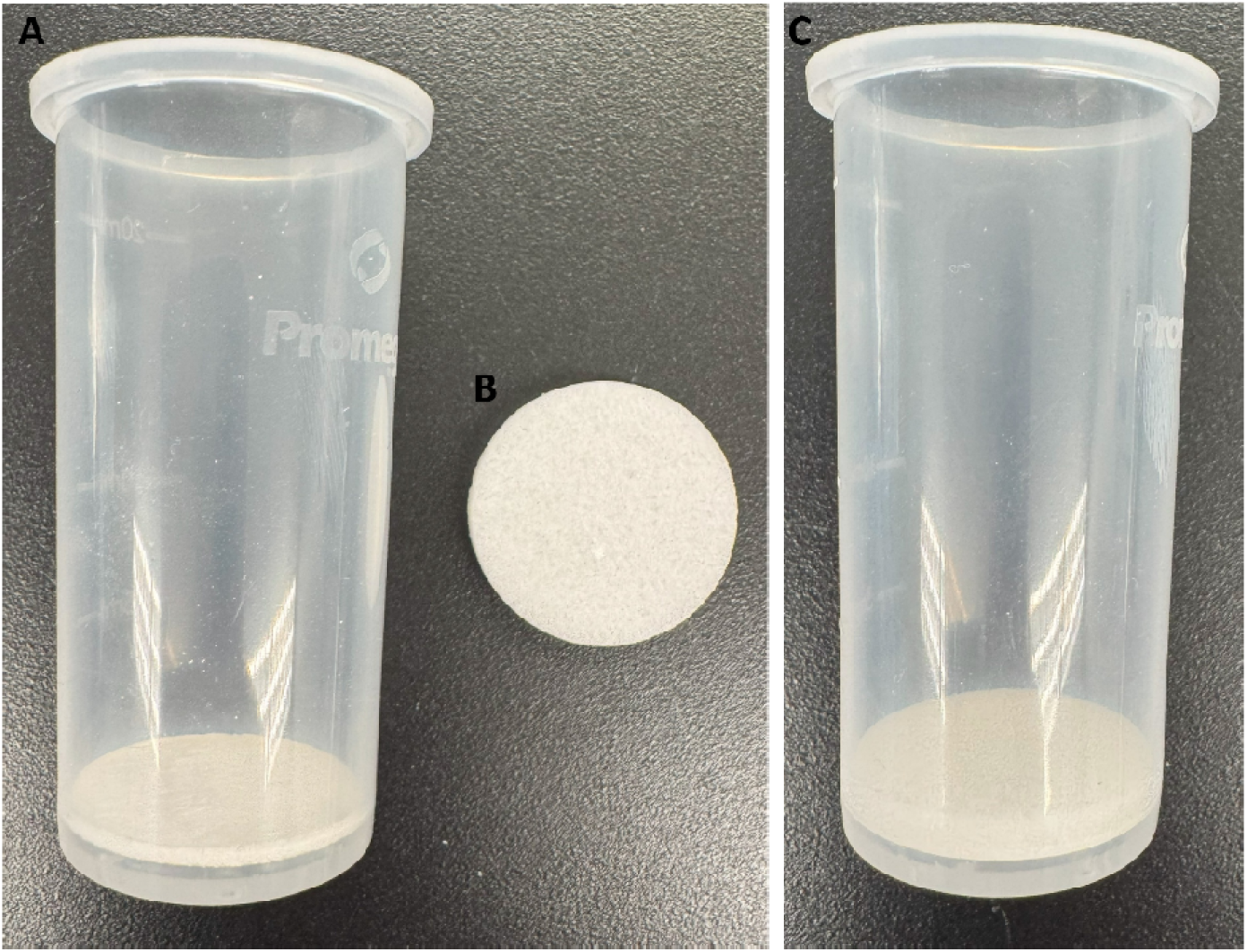
Development of PureBioX Xpurify Column. (A) The standard direct capture column from Promega Wizard® Enviro TNA Kit. (B) Hydrophobic filter (C) Assembled PureBioX Xpurify Direct Capture Column.

Two experiments were then performed to evaluate performance differences between the standard Promega column and the modified PureBioX Xpurify Column: (1) Maximum wastewater volume processed in 10 minutes (Figure 6) and (2) Processing speed and sensitivity using a standard 40 mL sample volume (Figure 7)

**Figure 6.**
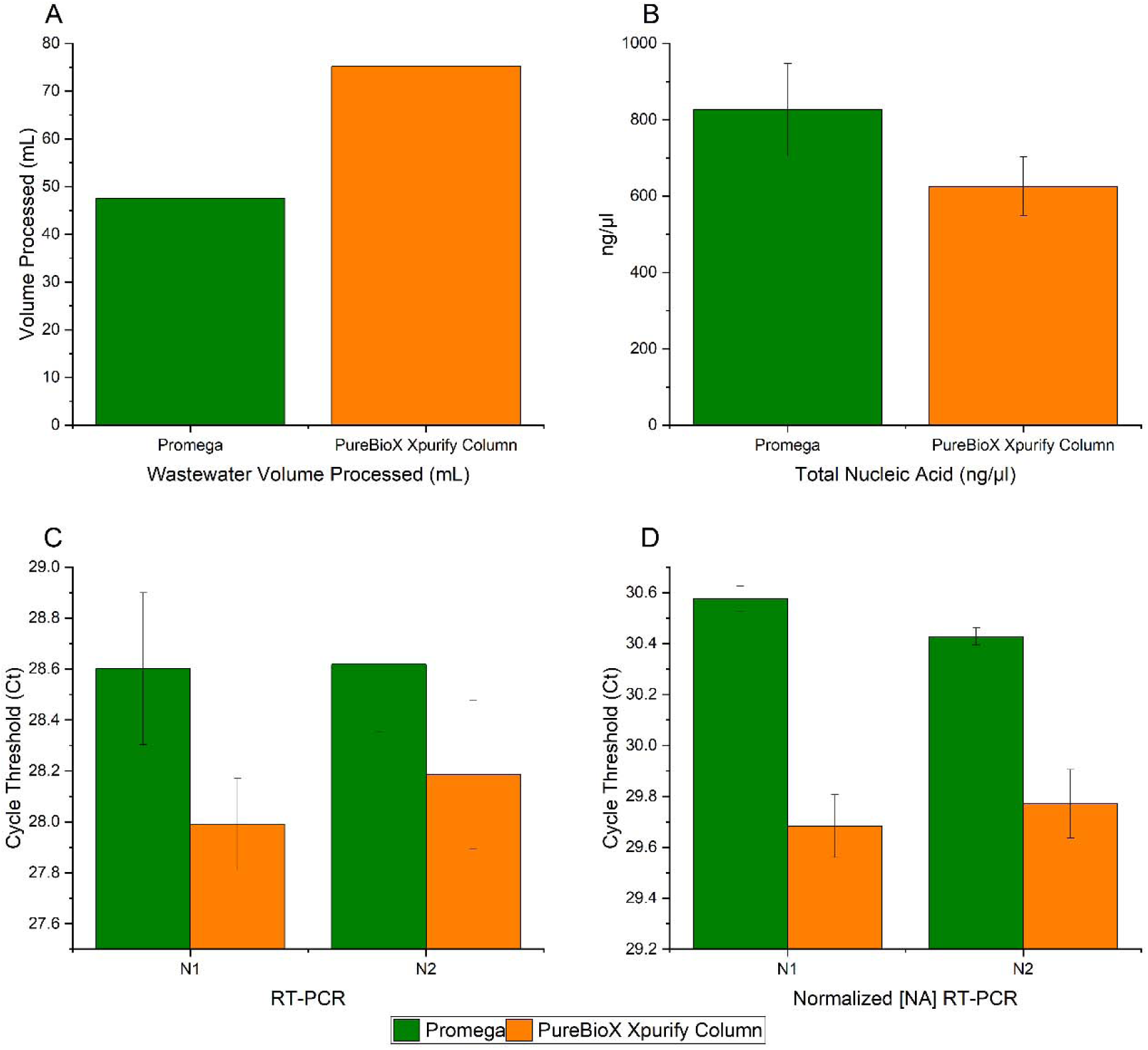
Comparison of the Promega Wizard® Enviro TNA Kit (green bars, *n* = 3) and the modified PureBioX Xpurify Column (orange bars, *n* = 3) for maximum wastewater processing in 10 minutes. (A) Volume of wastewater processed in 10 minutes. The PureBioX Xpurify Column processed 58.35% more volume (75.25 mL vs. 47.52 mL) than the Promega column (*p* = 0.044). (B) TNA yield (ng/µL) measured from a 40 µL elution using a NanoDrop One. Despite processing a larger volume, the PureBioX Xpurify Column yielded slightly lower TNA concentrations (625.7 vs. 825.8 ng/µL). (C) RT-PCR Ct values for SARS-CoV-2 nucleocapsid gene targets N1 and N2 using 5 µL of elution. The PureBioX Xpurify Column resulted in lower Ct values, indicating improved sensitivity (N1 *p* = 0.038, N2 *p* = 0.13). (D) RT-PCR Ct values using normalized input of 862 ng of TNA per reaction (based on lowest concentration sample). The PureBioX Xpurify Column demonstrated significantly lower Ct values, supporting increased viral RNA recovery and/or improved inhibitor removal (N1 *p* = 0.000, N2 *p* = 0.001).

**Figure 7.**
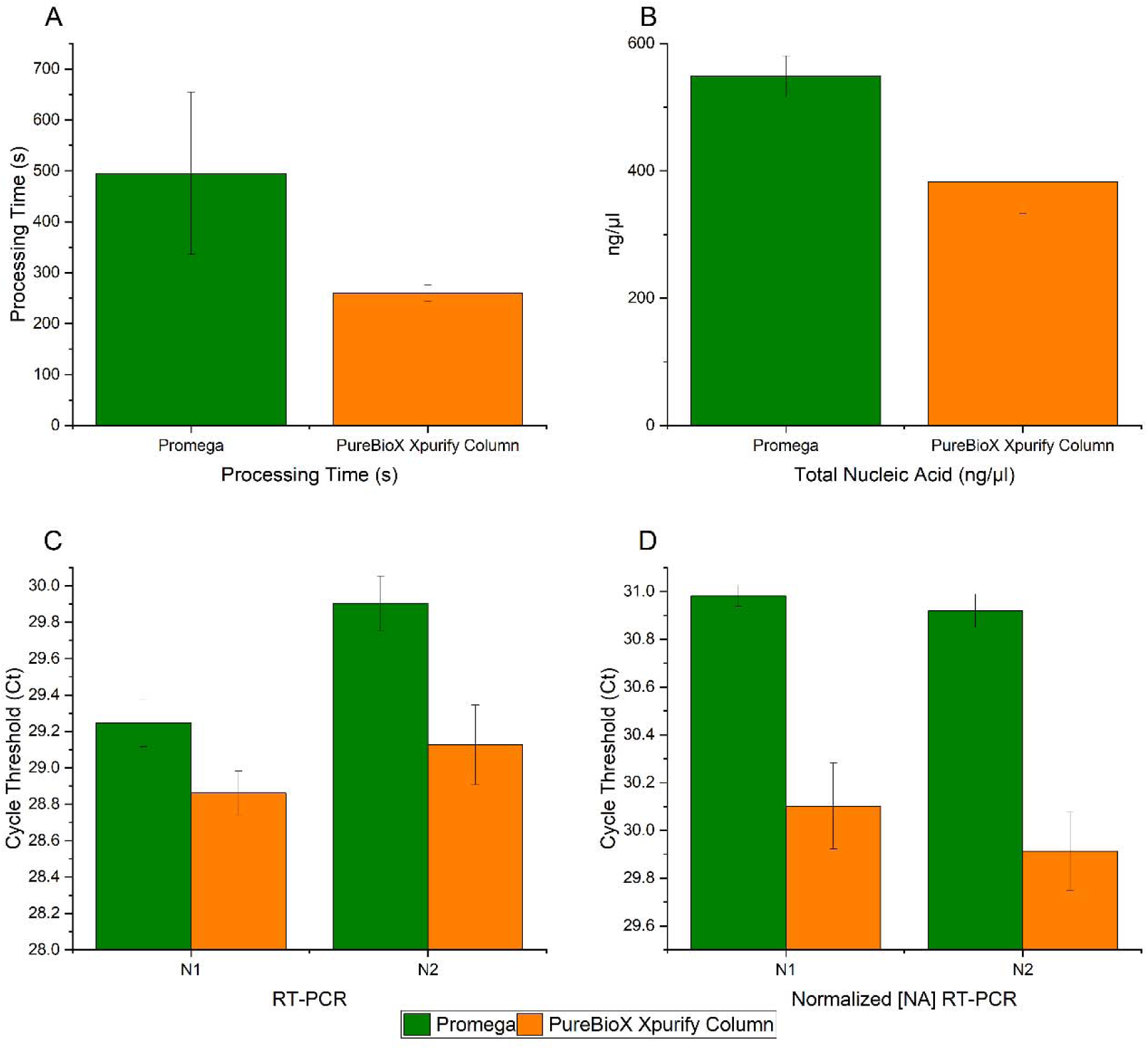
Comparison of the Promega Wizard® Enviro TNA Kit (green bars, *n* = 3) and the modified PureBioX Xpurify Column (orange bars, *n* = 3) for processing a fixed volume of 40 mL of wastewater. (A) Time required to process 40 mL of wastewater. The PureBioX Xpurify Column reduced processing time by 47.54% (259.67 seconds vs. 495.00 seconds) (*p* = 0.064). (B) TNA yield (ng/µL) from a 40 µL elution, measured via NanoDrop One. The PureBioX Xpurify Column yielded lower TNA concentrations (383.01 vs. 548.78 ng/µL, *p* = 0.008). (C) RT-PCR Ct values for SARS-CoV-2 nucleocapsid gene targets N1 and N2 using 5 µL of elution. Despite lower TNA yield, the PureBioX Xpurify Column produced lower Ct values, indicating improved sensitivity (N1 *p* = 0.019, N2 *p* = 0.007). (D) RT-PCR Ct values using normalized input of 185 ng TNA per reaction (ng input was based on the least concentrated sample). The PureBioX Xpurify Column yielded significantly lower Ct values, suggesting enhanced viral RNA recovery and/or improved inhibitor removal (N1 *p* = 0.001, N2 *p* = 0.0006).

In the first experiment, the PureBioX Xpurify Column processed 58.35% more wastewater in 10 minutes than the Promega column, handling an average of 75.25 mL compared to 47.52 mL (Figure 6A). In the second experiment, the time required to process 40 mL of wastewater was reduced by 47.54%, from 495.00 seconds using the Promega column to 259.67 seconds with the PureBioX Xpurify Column (Figure 7A).

Although the Promega column yielded higher total nucleic acid (TNA) concentrations in both experiments—825.8 ng/µL versus 625.7 ng/µL in experiment one, and 548.78 ng/µL versus 383.01 ng/µL in experiment two—RT-PCR analyses consistently showed that the PureBioX Xpurify Column produced lower Ct values for both the N1 and N2 targets of the SARS-CoV-2 nucleocapsid gene (experiment one: N1 *p* = 0.038, N2 *p* = 0.13; experiment two: N1 *p* = 0.019, N2 *p* = 0.007) (Figures 6B–C and 7B–C), indicating improved sensitivity for viral RNA detection.

Additionally, when RT-PCR reactions were normalized by total input nucleic acid (using the same nanogram amount per reaction) rather than the industry standard of 5 µL of elution per reaction, the PureBioX Xpurify Column showed a significant decrease in Ct values both when processing the maximum volume (N1 *p* = 0.000, N2 *p* = 0.001) (Figure 6D) and when processing the same 40 mL volume (N1 *p* = 0.001, N2 *p* = 0.001) (Figure 7D).

These results suggest that, despite a modest reduction in overall nucleic acid yield, the PureBioX Xpurify Column improves downstream detection sensitivity—potentially due to: (1) Enhanced viral RNA capture efficiency by the HP filter, (2) Reduction of PCR inhibitors in the final eluate, (3) Or a combination of both.

### Evaluation of Inhibitor Removal by the PureBioX Xpurify Column

To assess whether the increased sensitivity observed with the PureBioX Xpurify Column is due to enhanced removal of molecular inhibitors, an experiment was conducted using a controlled system. Specifically, 40 mL of deionized water—free of natural inhibitors—was spiked with nucleic acid previously purified from wastewater and processed using either the Promega column or the PureBioX Xpurify Column (Figure 8).

**Figure 8.**
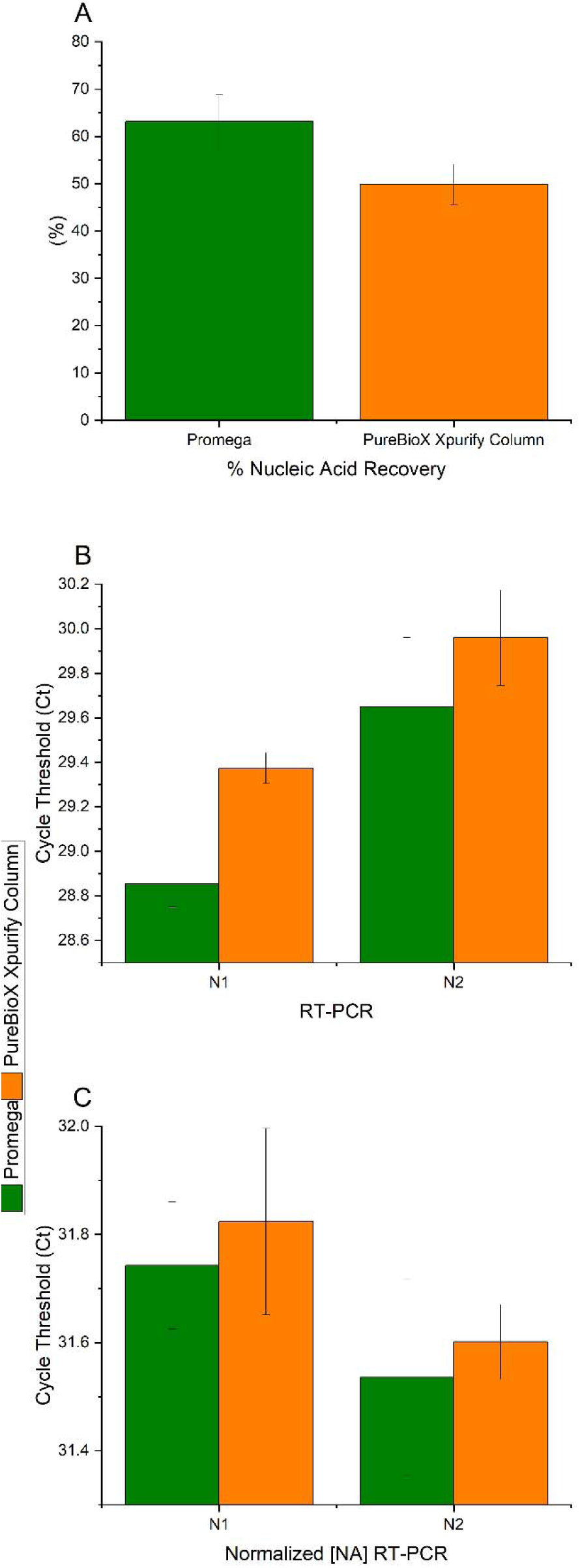
Processing of inhibitor-free Millipore water spiked with nucleic acid previously purified from wastewater, using Promega columns (green bars, *n* = 3) or PureBioX Xpurify Columns (orange bars, *n* = 3). (A) Total nucleic acid (TNA) yield (ng/µL) recovered during purification. The PureBioX Xpurify Column yielded statistically significantly less TNA than the Promega column (572.11 ng/µL vs. 724.11 ng/µL; *p* = 0.031). (B) Cycle threshold (Ct) values from RT-PCR targeting two regions of the SARS-CoV-2 nucleocapsid gene (N1 and N2), using 5 µL of unadjusted eluate as input. Higher Ct values for the PureBioX column reflect lower TNA input and the absence of inhibitors (N1 *p* = 0.002, N2 *p* = 0.228). (C) Ct values from RT-PCR using normalized nucleic acid input (515 ng per reaction, 1 µL of least concentrated sample), demonstrating no significant difference between columns (N1 *p* = 0.537, N2 *p* = 0.592). This indicates comparable amplification efficiency when input is controlled, supporting the conclusion that Ct differences under non-normalized conditions are due to recovery differences rather than inhibition.

Because inhibitor-free water was used, the expectation was that TNA yield would correlate directly with RT-PCR sensitivity: higher TNA recovery should result in lower Ct values. As shown in Figure 8A, the PureBioX Xpurify Column yielded lower TNA concentrations compared to the Promega column (572.11 ng/µL vs. 724.11 ng/µL; *p* = 0.031), consistent with reduced nucleic acid recovery due to the addition of the HP filter. Correspondingly, Ct values were higher for the PureBioX Xpurify Column (N1 *p* = 0.002, N2 *p* = 0.228) (Figure 8B), demonstrating that in the absence of inhibitors, sensitivity correlates with TNA yield.

However, when RT-PCR reactions were normalized for total nucleic acid input—ensuring the same nanogram quantity of starting material in each reaction—the Ct values between the two conditions were not statistically different (N1 *p* = 0.537, N2 *p* = 0.592). This confirms that the observed Ct differences under non-normalized conditions are driven by differences in recovery, not by amplification efficiency or inhibitor presence.

These findings contrast with results from wastewater samples (Figures 6 and 7), where the PureBioX Xpurify Column produced lower Ct values despite yielding less nucleic acid. This reversal strongly suggests that the HP filter improves sensitivity in complex matrices by reducing the carryover of RT-PCR inhibitors. In sum, these results support the conclusion that the enhanced detection sensitivity offered by the PureBioX Xpurify Column arises, at least in part, from its ability to reduce inhibitory substances in challenging environmental samples such as wastewater.

## Discussion

Wastewater-based epidemiology (WBE) is a critical tool for monitoring public health, offering near real-time surveillance of community-wide infectious disease trends, antimicrobial resistance, and other health indicators. As the global demand for actionable, high-quality WBE data grows, improvements in sample collection, nucleic acid extraction, and inhibitor management are essential to increase the accuracy, sensitivity, and utility of these efforts (1).

In this study, we demonstrate that simple but strategic column configurations can markedly improve direct capture columns performance for pathogen detection in wastewater. Specifically, we developed a novel stacked column incorporating a hydrophobic (HP) filter layer, herein referred to as the PureBioX Xpurify Column, with the dual goals of increasing filtration capacity and reducing molecular inhibitors that compromise downstream molecular assays.

Initial testing compared glass fiber filters to traditional silica membranes and found that the glass fiber matrix significantly improved total nucleic acid (TNA) yield. However, Ct values did not show a statistically significant difference despite higher yields. This discrepancy may be due to the heterogeneous and complex nature of wastewater, which contains a broad spectrum of inhibitory substances. Additionally, while the glass fibers may bind nucleic acids more effectively than silica, they may also allow more inhibitors to pass through, limiting their standalone utility.

Building upon this finding, we introduced a hydrophobic filter layer designed to sequester common inhibitors such as lipids, hydrocarbons, and detergents. This novel column yielded substantial improvements: the PureBioX Xpurify Column processed over 150% increase in wastewater in the same amount of time compared to the standard glass fiber-based column, and consistently produced lower Ct values despite yielding lower total nucleic acid in some conditions. This inverse relationship between TNA yield and RT-PCR sensitivity strongly suggests that the HP filter is effective at removing inhibitory substances, thereby enhancing molecular assay performance.

When tested in the context of a widely used commercial system—the Promega Wizard® Enviro TNA Kit—the addition of the HP layer significantly improved throughput, enabling the processing of larger wastewater volumes in shorter timeframes. Notably, in both maximum-volume and fixed-volume comparisons, the PureBioX Xpurify Column showed lower Ct values for SARS-CoV-2 targets, even when total nucleic acid yields were lower. This reinforces the hypothesis that reducing inhibitors is more impactful for RT-PCR sensitivity than increasing nucleic acid yield alone.

Importantly, we confirmed these effects using a controlled system in which purified nucleic acids were spiked into inhibitor-free water. Under these conditions, higher TNA recovery directly correlated with lower Ct values, and Ct differences between columns disappeared when reactions were normalized to contain equal amounts of nucleic acid. This provides strong evidence that in real wastewater matrices, the performance gains observed with the HP filter stem from its ability to reduce inhibitors, not differences in amplification efficiency or nucleic acid purity.

These findings have practical implications for public health agencies and laboratories engaged in wastewater monitoring. The modification described here is cost-effective, easy to implement, and compatible with existing workflows. It enables the processing of larger sample volumes and improves sensitivity—critical factors for detecting low-abundance targets during early outbreak detection or in populations with low infection prevalence.

While the current study demonstrates clear advantages of the PureBioX Xpurify Column, further research is warranted to optimize the filter design for other nucleic acid targets, sample types (e.g., stormwater, industrial effluent), and downstream applications such as metagenomic sequencing. Additionally, quantitative assessments of inhibitor removal—such as using external amplification controls or spectroscopic characterization of eluates—would further validate the mechanism of improvement.

Moreover, expanding these tests across varied geographic regions and wastewater compositions will help generalize the utility of this method. As WBE continues to scale globally, especially in low-resource settings, such adaptable and robust extraction enhancements could help democratize access to reliable wastewater surveillance.

In summary, the addition of a hydrophobic filter layer to standard direct capture columns significantly enhances wastewater processing by increasing throughput and improving sensitivity of RT-PCR detection. This simple yet effective column design reduces the burden of molecular inhibitors in complex environmental samples and enables more reliable pathogen surveillance. As public health decisions increasingly rely on wastewater data, innovations like the PureBioX Xpurify Column will be vital in making WBE more scalable, sensitive, and impactful.

## Data Availability

All data produced in the present study are available upon reasonable request to the authors

## Declarations

I, Lauren Aufdembrink, declare that all the above data was generated by authors of this paper or properly citied. The work provided here is the work of the authors and has not previously been published except where acknowledged. Lauren Aufdembrink wrote the manuscript, designed and performed the experiments, and processed the data. Akli Zarouri, Anil Meher, and Abdennour Abbas assisted in experimental design, data interpretation and manuscript editing.

## Acknowledgements

We would like to thanks the laboratory staff at the Metropolitan Wastewater Treatment Plant in St. Paul, MN for providing wastewater samples: Trent Staves, John Sipe, Bryan Henke and Steven Louwerse. This work was supported by a SBIR grant from the USDA (Award Number 2022-04393).

## Supplemental Figures

**Supplemental Figure 1.**
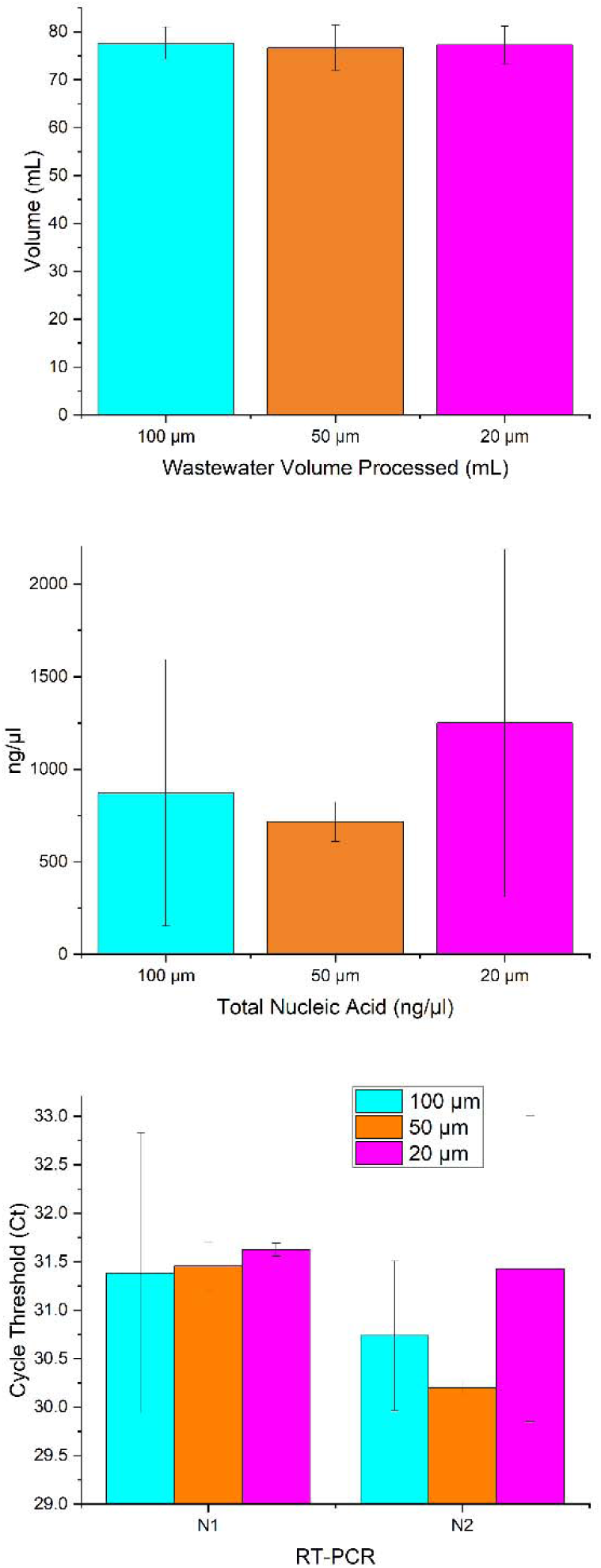
Evaluation of hydrophobic filter pore size in stacked column design. Columns were assembled using a hydrophobic filter placed above Cytiva glass fiber layers. Three pore sizes were tested: 100 µm (cyan bars, *n* = 3), 50 µm (orange bars, *n* = 3), and 20 µm (pink bars, *n* = 3). (A) Total volume of wastewater processed within 10 minutes. (B) Total nucleic acid (TNA) yield (ng/µL) from the final 40 µL elution, as measured by Nanodrop One. (C) RT-PCR cycle threshold (Ct) values for detection of SARS-CoV-2 nucleocapsid gene regions N1 and N2, using 5 µL of the final elution.

**Supplemental Figure 2.**
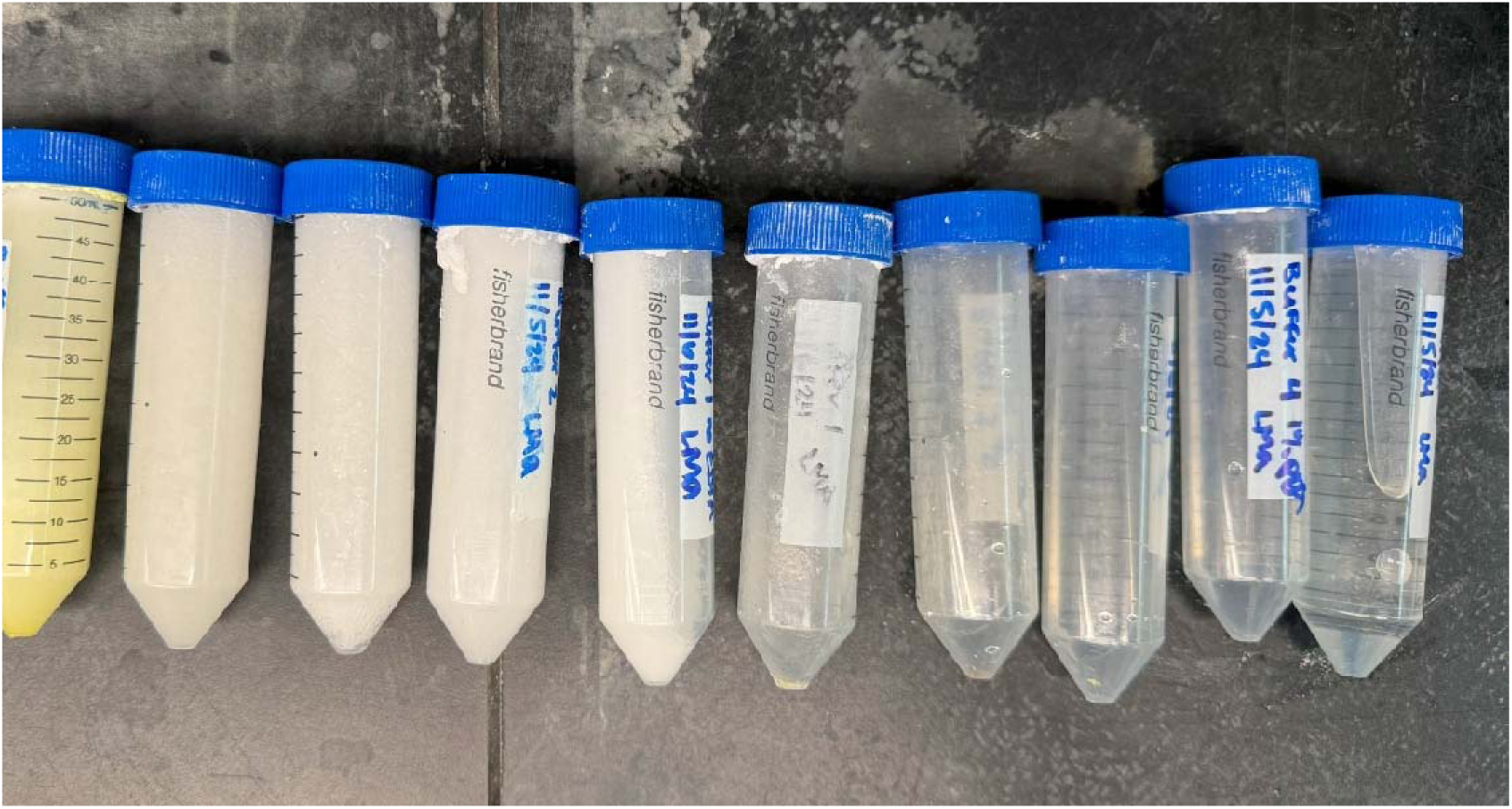
Image of lysis buffers tested. Buffers are at room temperature and all contain visible salt precipitation.

